# Resting-state functional MRI signal fluctuations are correlated with brain amyloid-β deposition

**DOI:** 10.1101/2021.04.22.21255924

**Authors:** Norman Scheel, Takashi Tarumi, Tsubasa Tomoto, C. Munro Cullum, Rong Zhang, David C. Zhu

## Abstract

Mounting evidence suggests that amyloid-β (Aβ) and vascular etiologies are intertwined in the pathogenesis of Alzheimer’s disease. Spontaneous fluctuations of the brain blood-oxygen-level-dependent (BOLD) signal, as measured by resting-state functional MRI (rs-fMRI), have been shown to be associated with neuronal activities as well as cerebrovascular hemodynamics. Nevertheless, it is unclear if rs-fMRI BOLD fluctuations are associated with brain Aβ deposition in individuals with an elevated risk of Alzheimer’s disease.

We recruited 33 patients with amnestic mild cognitive impairment who underwent rs-fMRI and positron emission tomography (PET). The Aβ standardized uptake value ratio (SUVR) was calculated with cortical white matter as the reference region to improve sensitivity for cortical Aβ quantification. We calculated the amplitudes of low-frequency fluctuations (ALFF) of local BOLD signals in the frequency band of 0.01-0.08 Hz. Applying physiological/vascular signal regression in stepwise increasing levels on the rs-fMRI data, we examined whether local correlations between ALFF and brain Aβ deposition were driven by vascular hemodynamics, spontaneous neuronal activities, or both.

We found that ALFF and Aβ SUVR were negatively correlated in brain regions involving the default-mode and visual networks, with peak correlation at the precuneus, and angular, lingual, and fusiform gyri. Regions with higher ALFF had less Aβ accumulation. The correlated cluster sizes in MNI space were reduced from 3018 mm^3^ with no physiological/vascular regression to 1072 mm^3^ with strong physiological/vascular regression, with mean cluster *r* values at approximately -0.47.

Results demonstrate that both vascular hemodynamics and neuronal activities, as reflected by BOLD fluctuations, are negatively associated with brain Aβ deposition. These findings further imply that local brain blood fluctuations due to either vascular hemodynamics or neuronal activities can affect Aβ homeostasis.

## Introduction

Alzheimer’s disease is a major neurodegenerative brain disorder and the most common type of dementia among older adults.^1^ Patients with Alzheimer’s disease exhibit memory loss, impaired decision-making capacity, disorientation, as well as changes in daily activity.^2^ These cognitive deficits and functional changes are accompanied and often preceded by brain pathophysiological changes including amyloid-β (Aβ) and tau protein depositions in the cerebral cortex, brain hypoperfusion, and neuronal degeneration.^3^ Mild cognitive impairment, particularly amnestic mild cognitive impairment, has been considered to represent a transitional phase between cognitive aging and Alzheimer’s disease, as many who develop amnestic mild cognitive impairment progress to Alzheimer’s disease.^4^

Two major hypotheses have been developed to understand the pathophysiological mechanisms of Alzheimer’s disease and to develop effective treatments. In the classic amyloid cascade hypothesis, the disruption of brain Aβ homeostasis has been proposed to be a primary driver of Alzheimer’s disease, leading to accumulations of Aβ plaques and neurofibrillary tangles, and consequently neurodegeneration and cognitive impairment.^5^ Conversely, the vascular hypothesis of Alzheimer’s disease proposes that cerebrovascular dysfunction is an important contributor to Alzheimer’s disease onset and progression.^6–8^ Mounting evidence indicates that Aβ and vascular etiology are intertwined in the pathogenesis of Alzheimer’s disease.^9,10^ In a recent study, we found that age-related carotid artery stiffness was associated positively with brain Aβ burden in patients with amnestic mild cognitive impairment, supporting the role of arterial aging in brain Aβ deposition.^11^ Recent animal and human studies also suggested that cerebral arterial oscillations and vasomotions influence brain Aβ clearance and deposition.^12–14^ Spontaneous cerebral blood flow (CBF) fluctuations are linked intrinsically to brain neuronal activities through neurovascular coupling.^15–17^ This phenomenon has been exploited by using resting-state functional MRI (rs-fMRI) to understand functional connectivities between brain regions and the disruption of functional connectivity in diseased states.^16,18–26^ Specifically, it has been proposed that spontaneous neuronal activities, which consume approximately 95% of the brain’s metabolism,^16^ are manifested as rs-fMRI blood-oxygen-level-dependent (BOLD) signal fluctuations.^16,17,27^

It has been widely recognized that spontaneous BOLD fluctuations are influenced by both systemic and cerebrovascular hemodynamics in addition to neuronal activities.^28–32^ In this regard, our previous work has shown that spontaneous fluctuations of BOLD signals and CBF velocity measured in the middle cerebral artery (upper-stream vascular signals) had similar spectral distributions and high correlation in the 0.01 - 0.08 Hz frequency band.^33^ These observations suggested that upper-stream cerebrovascular hemodynamic fluctuations can transmit downstream into the cerebral microcirculation and contribute importantly to regional BOLD fluctuations. On the other hand, local cerebral vasomotion, which has been linked to brain Aβ clearance, also may influence rs-fMRI BOLD fluctuations.^12^ A recent study also showed increased fMRI signal variability at cardiorespiratory frequencies in Alzheimer’s disease patients when compared to cognitively normal performing older adults.^34^ This study aimed to determine whether BOLD signal fluctuations as quantified by the amplitude of low-frequency fluctuations (ALFF) in the range of 0.01-0.08 Hz are related to brain Aβ deposition in patients with amnestic mild cognitive impairment.^4,35–38^ Furthermore, we examined whether the associations between local ALFF and brain Aβ depositions are influenced by the estimated upper-stream cerebrovascular hemodynamics, spontaneous neuronal activities, or both, using the well-established signal regression approaches.^39–41^ We hypothesized that local ALFF would be negatively correlated with brain Aβ deposition and that correlation strength between ALFF and Aβ deposition would decrease when the upper-stream vascular signals are regressed out from the raw BOLD signal under an assumption that upper-stream vascular effects drive brain Aβ clearance.

## Materials and methods

### Participants

Thirty-three amnestic mild cognitive impairment subjects (64.4 ± 6.4 years of age, 19 females) were recruited who represent a subgroup of participants enrolled in a proof of concept investigation aimed to identify effects of exercise training on neurocognitive function in amnestic mild cognitive impairment (ClinicalTrials.gov, NCT01146717).^42^ Data presented here were obtained at baseline before any intervention. The diagnosis of amnestic mild cognitive impairment was based on the Petersen criteria as modified by the Alzheimer’s Disease Neuroimaging Initiative (ADNI) project (http://adni-info.org).^43^ Diagnostic assessments included the Clinical Dementia Rating (CDR) scale,^44^ the Mini-Mental State Examination (MMSE),^45^ and the Wechsler Memory Scale-Revised Logical Memory (LM) immediate and delayed recall trials.^46^ Heart rate (HR) and brachial cuff blood pressure (BP) were measured >3 times with an ECG-gated electro-sphygmomanometer (Suntech, Morrisville, NC, USA). Obtained values were averaged to obtain HR, systolic (SBP), and diastolic BP (DBP). Participants with major psychiatric disorders, major or unstable medical conditions, uncontrolled hypertension, diabetes mellitus, or chronic inflammatory diseases were excluded, as were participants with a cardiac pacemaker or any metal plates or pins in their body preventing them from undergoing MRI scanning (detailed inclusion and exclusion criteria are provided in ClinicalTrials.gov NCT01146717). All participants gave informed consent. This study was approved by the Institutional Review Boards of the University of Texas Southwestern Medical Center and Texas Health Presbyterian Hospital of Dallas and performed by the guidelines of the Declaration of Helsinki and Belmont Report.

### MRI Measurements

Rs-fMRI data were collected on a Philips Achieva 3T scanner (Philips Healthcare, Best, the Netherlands) under an “eye-closed” condition with the following parameters: gradient recalled echo planner imaging (EPI), 29 contiguous 5-mm axial slices, 30-ms time of echo (TE), 1500-ms time of repetition (TR), 60° flip angle, a 24-cm field of view, 80 × 80 matrix size and 200 time-points. High-resolution T_1_-weighted 3D MPRAGE (Magnetization-Prepared Rapid Acquisition Gradient-Echo) images were also collected to cover the whole brain with the following parameters: TE/TR = 3.7/8.1 ms, flip angle = 12°, field of view = 256 mm × 204 mm, 160 1-mm slices, resolution = 1 mm × 1 mm × 1 mm, SENSE factor = 2 and scan duration = 4 minutes.

### PET image acquisition

After an intravenous bolus injection of 10 mCi ^18^F-florbetapir (also called AV45), participants were positioned in a Siemens (Munich, Germany) ECAT HR PET scanner for data acquisition, using laser guidance for precise head positioning. Velcro straps and foam wedges were used to secure the participant’s head. To ensure the brain was completely in the field of view and absent of rotation in either the transverse or sagittal planes, a 2-minute scout scan was acquired. At 50 minutes post-injection, 2 frames of 5-minute PET emission scan and a 7-minute transmission scan were acquired in 3D mode using the following parameters: matrix size = 128 × 128, resolution = 5 mm × 5 mm, slice thickness = 2.42 mm, and field of view = 58.3 cm. Emission images were processed by iterative reconstruction, 4 iterations, and 16 subsets with a 3-mm full width at half maximum (FWHM) ramp filter. The transmission image was reconstructed using back-projection and a 6-mm FWHM Gaussian filter for attenuation correction ^47,48^.

### RS-fMRI BOLD signal fluctuation quantification

The quantification of rs-fMRI BOLD local signal fluctuation using ALFF was first proposed by Zang et al..^38^ It was first used to investigate regional rs-fMRI signal fluctuations in attention deficit hyperactivity disorder. To calculate ALFF, rs-fMRI recordings are first pre-processed according to current standards, including slice-timing correction, motion correction, and spatial blurring (different strategies used here are detailed later). Importantly, no temporal filtering is applied before ALFF calculation. Fast Fourier transformation (FFT) is carried out on each preprocessed voxel time course. The ALFF index is calculated as two times the amplitude of the FFT output, divided by the sample length (number of data points in the time course), then averaged across the specified frequency band of interest.

A frequency band of 0.01 - 0.08 Hz was used in this study. Figure 1(a) shows a typical pre-processed rs-fMRI time course at a brain voxel. Figure 1(b) shows the corresponding Fourier spectrum and the frequency band for ALFF computation. ALFF at each voxel is subsequently normalized by the global mean ALFF from all voxels within the brain to generate a standardized ALFF index (sALFF) that allows comparisons across participants. ALFF computations were carried out using the ALFF function included in the DPARSFA toolbox from the “DPABI V4.3_200401” software package^49,50^ in MATLAB R2019a (MathWorks, Natick, MA, USA).

**Figure 1.**
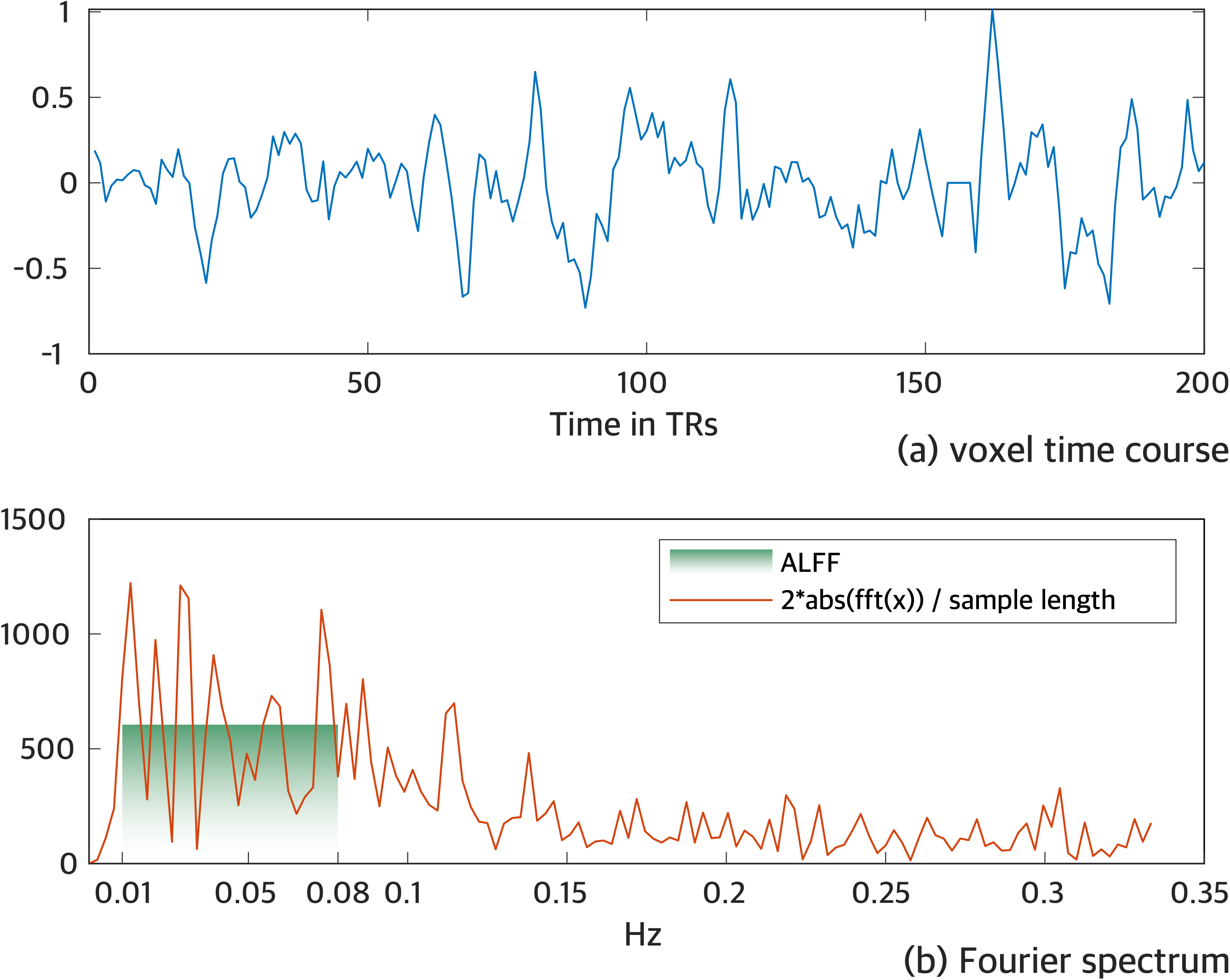
Time series and spectral analysis of spontaneous BOLD signal fluctuations. **(a)** time course at a voxel in the occipital region of one study participant; **(b)** corresponding Fourier spectrum. Across the applied frequency band of 0.01-0.08 Hz, the amplitude of low-frequency fluctuations (ALFF) yields 605.7 as of this voxel’s fluctuation amplitude (plotted in the green shade).

### Anatomical image pre-processing and MNI-registration

One common challenge in brain imaging studies of older adults is the high degree of anatomical variability due to atrophy, leading to difficulties in performing group-wise image analyses. FreeSurfer^51^, an automated data processing software for cortical surface reconstruction and anatomical segmentation of brain MRI scans, provides a robust brain segmentation in comparable study cohorts, as demonstrated in prior studies.^26^ The high-resolution T_1_-weighted volumetric MR images of each participant were processed using the default FreeSurfer “recon-all” pipeline. This pipeline creates a robust non-linear transformation matrix that warps images from a participant’s native space to MNI standard space, according to the MNI305 template resolution (1 mm × 1 mm × 1 mm with a matrix size of 256 × 256 × 256). This transformation matrix was applied to warp the results of the sALFF and AV45-SUVR calculations to the MNI305 template. While the transformation to the standard template performed well overall for brains with atrophy, artifacts in subcortical regions were noticeable due to non-linear stretching, especially for brains with enlarged ventricles. These artifacts generated spatial noise in our data analyses. Subsequently, for computational efficiency, all images in MNI305 template resolution were resampled to fit the MNI152 template with a resolution of 2 mm × 2 mm × 2 mm at a matrix size of 91 × 109 × 91.

### fMRI preprocessing and physiological/vascular signal regression

Resting-state fMRI pre-processing was first carried using AFNI software ^52^ in native space. The “afni_proc.py” routine in AFNI was used to generate the script to pre-process the rs-fMRI data. For each participant, any signal spikes in the signal intensity time courses were first detected and removed. The acquisition timing difference was then corrected for different slice locations. With the third functional volume as the reference, rigid-body motion correction was carried out in three translational and three rotational directions. The displacements due to motion in these six directions as well as the corresponding motion derivatives at each time point were estimated and then modeled as motion regressors. The data points with excess motion (normalized motion derivative□>□0.5 or voxel outliers□>□10%) were identified to generate another motion regressor. For each participant, spatial blurring with a full width half maximum (FWHM) of 4 mm was used to reduce random noise. At each voxel, motion-related signal changes, baseline, linear, 2nd-order, and 3rd-order system-induced signal trends were modeled as linear regressors. The “3dDeconvolve” function in AFNI was applied to remove the motion and system introduced noises from each voxel’s time course, as modeled in the regressors described above. Up to this step, physiological signals, likely due to upper-stream vascular effects, have not yet been regressed from the signal. The resulting fMRI dataset up to this step will be called “No Physiological Regression” to emphasize that no attempt has been made to remove upper-stream vascular effects.

The standardized amplitude of the BOLD signal fluctuations (sALFF) within the frequency range of 0.01 Hz to 0.08 Hz was then calculated for each voxel within the brain. For group analyses, subject-space sALFF maps were non-linearly transformed into MNI305 standard space, using the transformation matrix created by the FreeSurfer processing pipeline as discussed above. MNI305 sALFF images were subsequently resampled to fit the MNI152 standard template for improved computational efficiency. The correlation between sALFF and brain amyloid β accumulation, calculated as AV45 PET amyloid β standardized uptake value ratio (SUVR), was then calculated in MNI152 standard space as discussed further in the following section. To understand the upper-stream physiological/vascular effects, the “no physiological regression” dataset was further processed with three progressively more aggressive physiological signal regression procedures. For each procedure, sALFF was calculated again followed by the aforementioned standard-space transformations and correlation analyses. The three regression procedures, in addition to the “no physiological regression”, are listed below:

1. “No physiological regression:” as described above, this procedure has no special treatment to remove upper-stream physiological/vascular effects before sALFF computation.
2. “WM/CSF regression:” White matter (WM) and cerebrospinal fluid (CSF) do not contain neuronal brain activity. Consequently, fluctuations of mean BOLD signal time courses at these regions can be assumed to mostly represent upper-stream vascular effects.^53^ Hence, CSF and WM signals were modeled and used as additional regressors in the “3dDeconvolve” step to reduce the upper-stream vascular influence, before sALFF computation.
3. “WM/CSF/GS regression:” The mean global signal (GS) was assumed to be primarily driven by an overall upper-stream vascular effect and added as an additional regressor to this “WM/CSF regression” procedure for further physiological signal removal before sALFF computation. With this procedure, the resulting signal time course should closely reflect neuronal brain activity.^53^ While global signal removal can potentially remove some neuronal signals from fMRI data, current research suggests that the upper-stream vascular components most likely dominate the global signal.^54– 56^ Recently, Xifra-Porxas and colleagues reported that physiological signals accounted for an *r* of about 0.6 within the global signal.^57^ Here, global signal regression was used to further investigate the effect of the upper-stream vascular signals on Aβ deposition in the brain and its influence in this cascaded setup.
4. “Aggressive AROMA:” A rs-fMRI noise removing technique called “ICA-AROMA” was developed by Pruim and colleagues^39^ to aggressively remove motion artifacts and physiological noise. The authors demonstrated that this technique could effectively identify motion and physiological signals as independent components and then remove them from rs-fMRI data.^58^ “ICA-AROMA” was implemented using FSL’s MELODIC routine ^59^. In this technique, single-subject spatial independent component analysis (ICA) is performed on a motion-corrected whole-brain rs-fMRI dataset in native space. Using a pre-trained classifier, the resulting components are sorted into noise and non-noise components. The time courses of components identified as noise are subsequently regressed from the data using FSL’s “regfilt” function, where partial regression is called “non-aggressive AROMA,” and full regression is called “aggressive AROMA.” The “aggressive AROMA” approach was employed here to maximize the effect of removing upper-stream vascular signals.

The overall signal processing steps described above are also presented in supplementary Figure S1.

### PET image processing

Using “flirt” from the FSL software package,^59^ a six-degree linear transformation was applied to align PET AV45 SUVR images to the 3D MPRAGE anatomical MRI images. All alignments were visually inspected and improved with manual adjustments if necessary.

Using the segmentation of the high-resolution 3D MPRAGE image from the FreeSurfer analysis, white-matter (WM) and cerebellar regions were isolated. An additional erosion of the white-matter map, using FSL’s “fslmaths” with a box kernel of 4 mm × 4 mm × 4 mm ensured that only white-matter regions were obtained. The AV45 uptake images were then normalized by the mean uptake at the white matter and also the cerebellar regions to generate two versions of SUVR images. Previous studies have shown that SUVR calculation using WM signal can improve discriminatory power for detecting Alzheimer’s disease and mild cognitive impairment.^60^ In the following results, SUVR will be reported for WM normalization, yet posthoc analyses with the AV45-SUVR images normalized by cerebellar signal were also carried out to confirm observations. Congruent to the spatial normalization of sALFF maps, AV45-SUVR images were non-linearly transformed to match the MNI305 standard template, using the transformation matrix created by the FreeSurfer processing pipeline. AV45-SUVR images in MNI305 template space were subsequently resampled to fit the MNI152 standard template for better computational efficiency.

## Statistical analysis

Correlations between sALFF and AV45-SUVR across participants were carried out for each brain voxel in MNI152 template space. Monte-Carlo simulations, using 10,000 random permutations for each correlation value *r* were applied to approximate the corresponding *p* values, through constructing voxel-specific empirical cumulative distributions.^61^ These computations were realized in MATLAB R2019a (MathWorks, Natick, MA, USA). Cluster-level multiple-comparison corrections were computed using AFNI’s^52^ “3dClustSim” Monte-Carlo simulation software. In these simulations, we estimated the minimum cluster sizes required to achieve a corrected *p* ≤ 0.05 at a voxel-level *p* < 0.001. As the estimated minimum cluster size is highly dependent on data spatial smoothness, which in turn depends on data preprocessing, cluster size thresholds were estimated independently for each physiological/vascular signal removal procedure, described earlier. Autocorrelation function parameters (-acf) employed by “3dClustSim” were estimated using “3dFWHMx” (AFNI) on the preprocessed fMRI data (MNI transformed and resampled to 91 x 109 x 91 isotropic 2-mm voxels). This procedure yields mean parameters (across participants) for each of the four different autocorrelation functions, specific to each fMRI preprocessing procedure. We also estimated the spatial autocorrelation of the AV45-SUVR maps using “3dFWHMx”. Our pre-processed fMRI images showed less auto-correlation than the AV45-SUVR maps. To be conservative, we used the autocorrelation estimates of the fMRI images to calculate the required cluster sizes to achieve statistical significance. Subsequently, “3dClusterize” (AFNI) was used to generate correlation cluster maps that survive the estimated cluster correction thresholds for each preprocessing procedure.

The preprocessing-specific significant clusters were then used to assess whether the correlations between BOLD signal fluctuations and brain amyloid deposition were rather influenced by upper-stream physiological/vascular effects or neuronal activations. For this purpose, we illustrated the distributions of the Fisher *z* transformed *r* values of the AV45-SUVR-sALFF correlations, using the voxel populations identified as significant clusters resulting from the analyses based on the “no physiological regression” and the “aggressive AROMA” procedures.

## Data availability

The raw data were generated at the University of Texas Southwestern Medical Center and UTSW and Institute for Exercise and Environmental Medicine Texas Health Presbyterian Hospital, Dallas, TX, USA. Derived data supporting the findings of this study are available from the corresponding authors upon reasonable request.

## Results

Table 1 shows the participant characteristics. Overall, our amnestic mild cognitive impairment patients were well educated and showed early amnestic mild cognitive impairment symptoms, as shown by CDR scores of 0.5, a mean MMSE score of 29.5 ± 0.8, and mean LM immediate recall of 11.4 ± 2.3 and delayed recall of 9.7 ± 1.8. These older adults had an average heart rate of 62 bpm and mean systolic blood pressure of 118 mmHg but included hypertension (≥140 mmHg). The presence of apolipoprotein E4 (APOE4) was assessed in 28 participants and 25% were APOE4 positive. The participants had a mean cortical AV45-SUVR of 0.57 ± 0.05 when normalized with the white matter as a reference and 1.15 ± 0.08 with the whole cerebellum as a reference.

**Table 1.** Participant characteristics.

|  | Mean | ± | SD | Minimum | Maximum |
| --- | --- | --- | --- | --- | --- |
| Men/Women (n) |  |  |  | 14/19 |  |
| Race (n) |  |  |  | 29 Caucasian, 4 African American |  |
| APOE4 carrier*, n (%) |  |  |  | 7 (25%) |  |
| Age (years) | 64 | ± | 7 | 55 | 78 |
| Height (cm) | 169 | ± | 9 | 143 | 184 |
| Body mass (kg) | 79 | ± | 14 | 58 | 117 |
| Body mass index (kg/m<sup>2</sup>) | 28 | ± | 4 | 21 | 36 |
| Education (years) | 16 | ± | 2 | 12 | 18 |
| Clinical Dementia Rating (points) |  |  |  | 0.5 |  |
| MMSE total | 29.5 | ± | 0.8 | 27 | 30 |
| Logical Memory immediate recall | 11.4 | ± | 2.3 | 5 | 16 |
| Logical Memory delayed recall | 9.7 | ± | 1.8 | 6 | 15 |
| Heart rate (bpm) | 62 | ± | 9 | 44 | 81 |
| Systolic blood pressure (mmHg) | 118 | ± | 18 | 85 | 152 |
| Diastolic blood pressure (mmHg) | 70 | ± | 8 | 53 | 87 |

\*Apolipoprotein E4 (APOE4) was assessed in 28 patients.

Figures 2 and 3 show graphical representations of select sagittal and axial slices of mean AV45-SUVR, mean sALFF for each preprocessing procedure, and the significant correlation clusters between them. Table 2 provides a detailed list of significant correlation clusters with associated brain regions for each preprocessing procedure. Using the “no physiological regression” procedure, cluster level significance was estimated with voxel-level *p* < 0.001 and a corrected *p* ≤ 0.05, yielding a minimum cluster size of 493 voxels (2-mm isotropic). Using this threshold, four significant negatively correlated clusters between sALFF and AV45-SUVR were found spanning across the lingual gyrus, precuneus, cuneus, and fusiform gyrus, with the most prominent cluster reaching a size of approximately 3018 mm^3^ (see Table 2, Figures 2b & 3b).

**Table 2.**
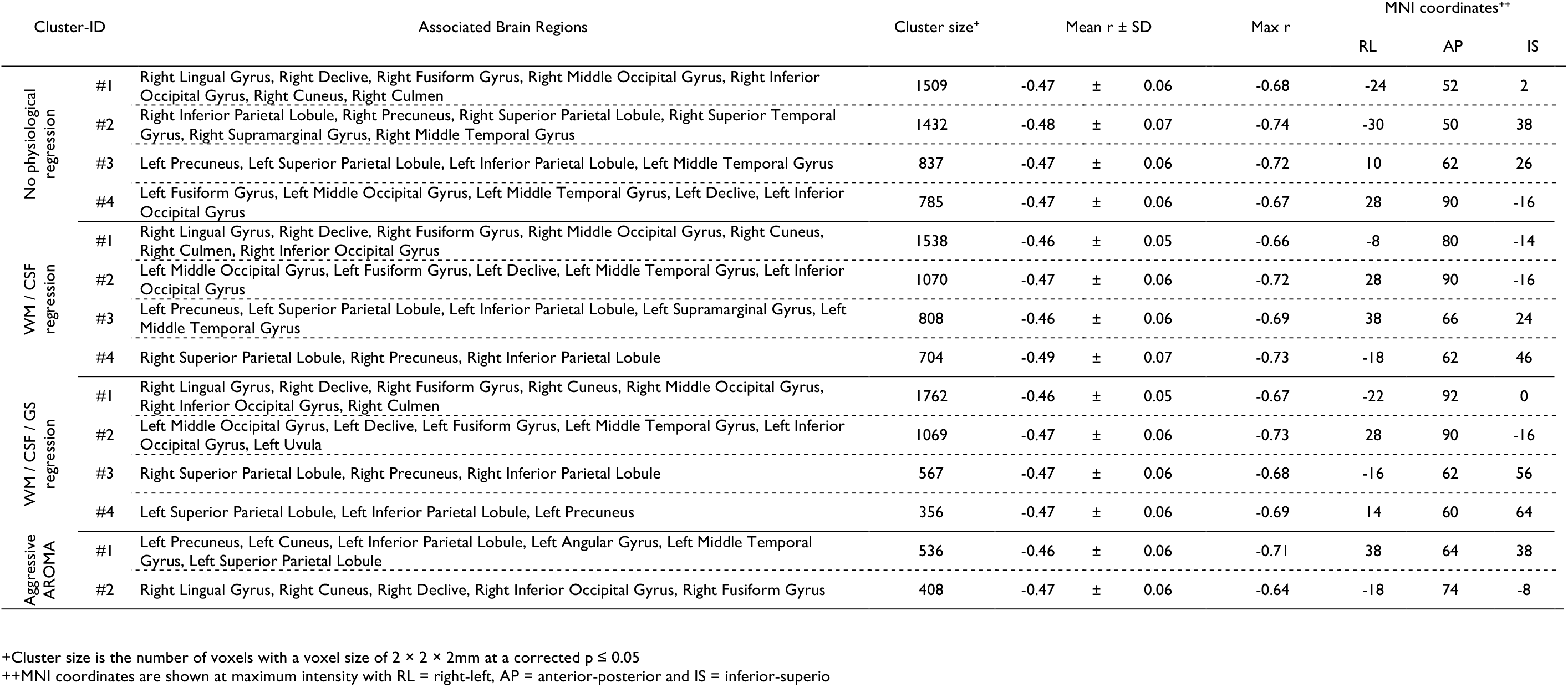
Correlation cluster analysis of sALFF and AV45-SUVR.

**Figure 2.**
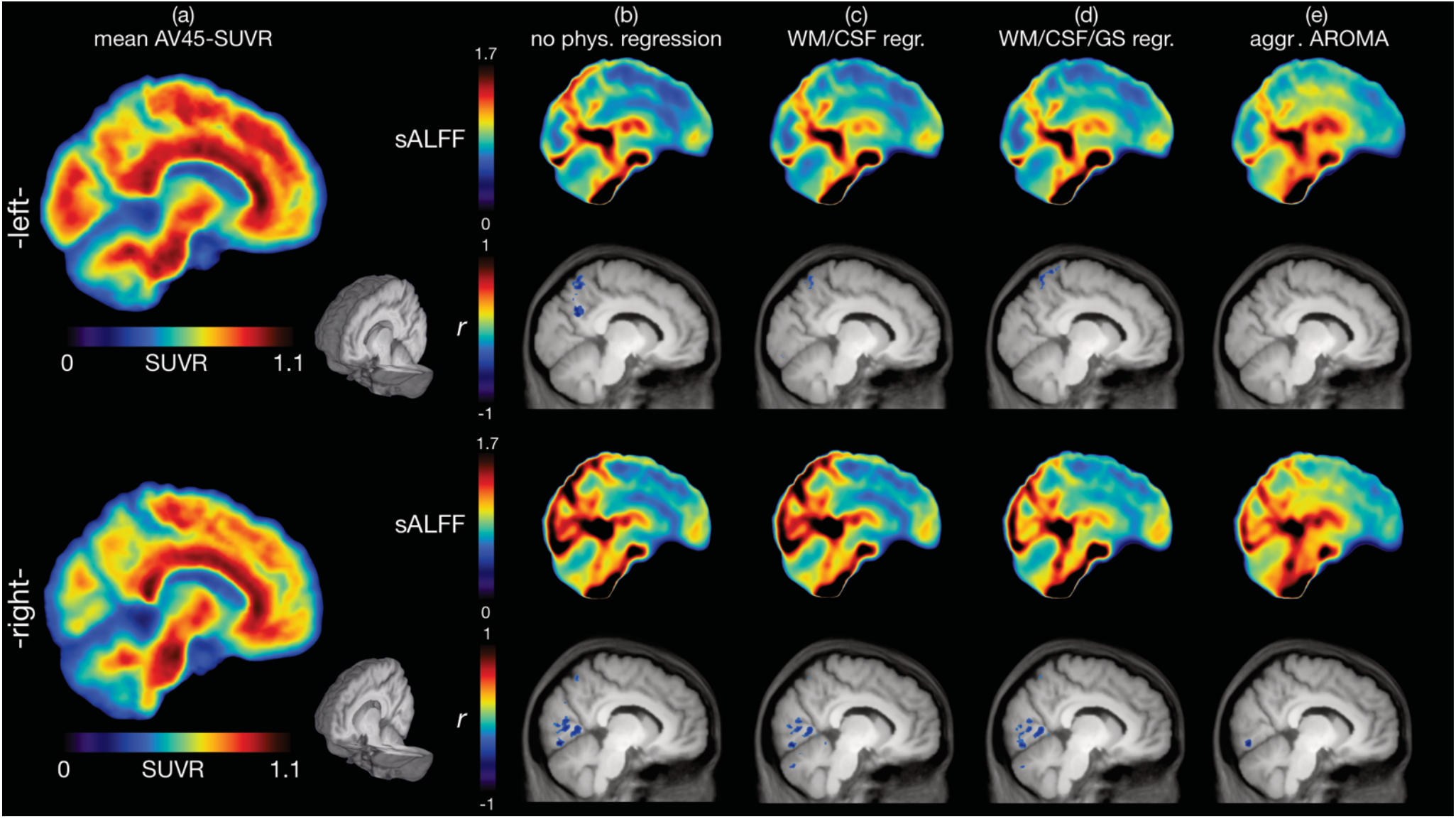
Voxel-wise correlation analysis between brain amyloid-β deposition and the amplitude of low-frequency BOLD signal fluctuations (sALFF) - sagittal. Images represent slices near the mid-sagittal region (right and left 9.25mm in MNI space). **(a)** mean AV45-SUVR (standardized uptake value ratio with the white matter as a scaling reference) color maps of 33 amnestic mild cognitive impairment participants are shown along with the corresponding mean sALFF (standardized amplitude of low-frequency fluctuations) color maps after the physiological signal cleaning procedures of **(b)** “no physiological regression”, **(c)** “WM/CSF regression”, **(d)** “WM/CSF/GS regression” and **(e)** “aggressive AROMA”. Significant correlation clusters between AV45-SUVR and sALFF for each regression procedure (b)-(e) are shown in the row below the mean sALFF images. Clusters are depicted in blue, as r values are negative (see Table 2 for detailed statistical analysis data). AV45-SUVR maps are nearly identical if the cerebellum, instead of white matter, is used as a reference, except the color scale range becomes 0 to 2.

**Figure 3.**
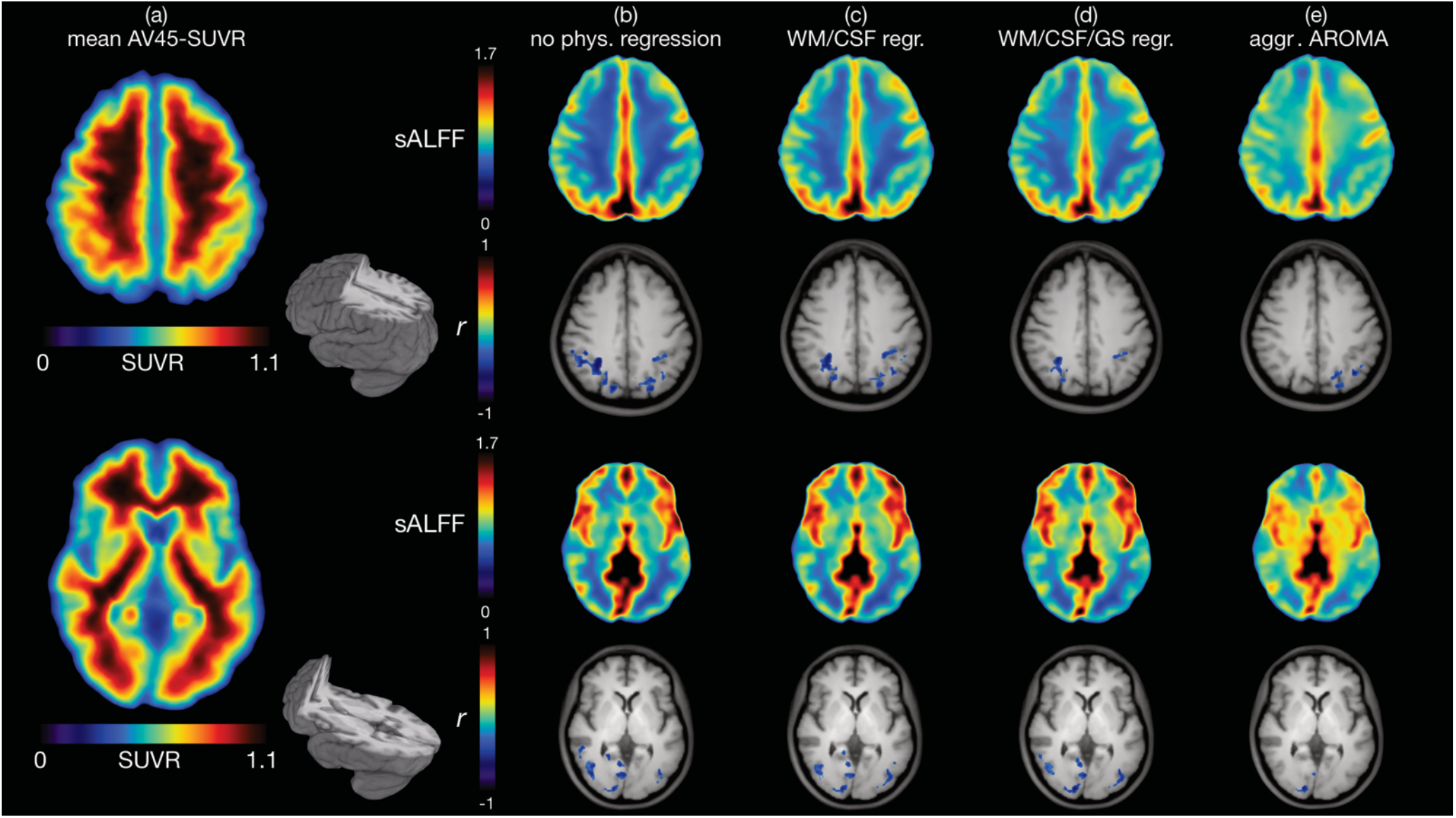
Voxel-wise correlation analysis between brain amyloid-β deposition and the amplitude of low-frequency BOLD signal fluctuations (sALFF) - axial. Images represent an axial slice (top half) above the corpus callosum (42mm in inferior-superior plane MNI) and an axial slice (bottom half) across the thalamus (0.75mm in inferior-superior plane MNI). **(a)** mean AV45-SUVR (standardized uptake value ratio with white matter scaling) color maps of 33 amnestic mild cognitive impairment participants are shown, along with the corresponding mean sALFF (standardized amplitude of low-frequency fluctuations) color maps after the physiological signal cleaning procedures of **(b)** “no physiological regression”, **(c)** “WM/CSF regression”, **(d)** “WM/CSF/GS regression”, and **(e)** “aggressive AROMA”. Significant correlation clusters between AV45-SUVR and sALFF for each regression procedure (b)-(e) are shown in the row below the mean sALFF images. Clusters are depicted in blue, as r values are negative (see Table 2 for detailed statistical analysis data). AV45-SUVR maps are nearly identical if the cerebellum, instead of white matter, is used as a reference, except the color scale range becomes 0 to 2.

After applying the “WM/CSF regression” procedure using the same statistical criteria as above, the corrected *p* ≤ 0.05 was achieved with a minimum cluster size of 434 voxels. At this threshold, four clusters were identified as significant. The significant brain regions are consistent with the “no physiological regression” methods, spanning the lingual gyrus, fusiform gyri, cuneus, and precuneus, yet at slightly smaller cluster sizes for all except the lingual gyrus cluster which slightly increases in size (see Table 2, Figures 2c & 3c).

After applying the “WM/CSF/GS regression” procedure using the same statistical criteria as above, the corrected *p* ≤ 0.05 was achieved with a minimum cluster size of 344 voxels. Again, four clusters were identified as significant at brain regions consistent with the prior procedures. While the cluster at the right lingual gyrus, fusiform gyrus, and cuneus increases in size once more, clusters at regions involving precuneus and left fusiform gyrus show smaller cluster sizes (see Table 2, Figures 2d & 3d).

After applying the “aggressive AROMA” procedure using the same statistical criteria as above, the corrected *p* ≤ 0.05 was achieved with a minimum cluster size of 384 voxels. Two clusters spanning precuneus, cuneus, angular gyrus, fusiform gyrus, and lingual gyrus were identified as significant, with the most prominent cluster reaching a size of approximately 1072 mm^3^ (see Table 2, Figures 2e & 3e).

Overall, significant negative correlations were found between sALFF and AV45-SUVR with all four regression procedures, decreasing in cluster sizes in a stepwise manner based on the aggressiveness of physiological/vascular signal removal. Scatterplots, demonstrating the negative correlation between the mean sALFF and mean AV45-SUVR at the location of the most significant cluster, found using the “no physiological regression” procedure, are depicted in Figure 4, across the four regression procedures. The corresponding scatterplots for all significant voxel populations across all regression procedures are presented in Supplementary Figures S2-S5.

**Figure 4.**
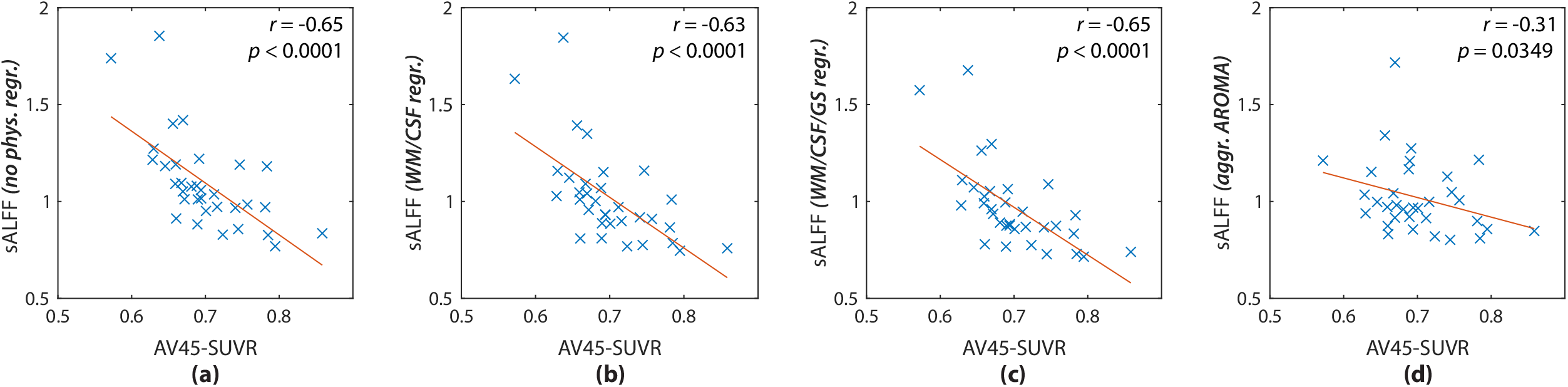
Correlation of brain amyloid-β deposition and the amplitude of low-frequency BOLD signal fluctuations (sALFF) for “No physiological regression” cluster #1 (Table 2). Depicted are scatterplots for each physiological/vascular regression procedure, left to right: **(a)** “no physiological regression”, **(b)** “WM/CSF regression”, **(c)** “WM/CSF/GS regression”, and **(e)** “aggressive AROMA”. Blue crosses represent the 33 subjects, using mean sALFF and mean AV45 within this cluster. The plots show reduced correlation strength from (a) to (d). For scatterplots of the remaining significant clusters see Supplementary Figures 2-5.

To further understand the effect of the different physiological/vascular signal regression procedures on sALFF and AV45-SUVR correlations, the Fisher z transformed *r*-value distributions at significant voxel locations were illustrated in Figure 5. Using all significant voxel locations for either the “no physiological regression” procedure (4563 voxels) or the “aggressive AROMA” procedure (944 voxels), *r*-value distributions are depicted for each regression procedure at these specific voxel locations. Figure 5 (a) shows the violin plots of the *r*-value distributions of significant voxels from the no physiological/vascular regression procedure with mean and standard deviations as follows: - 0.47 ± 0.06 (no physiologic regression), -0.45 ± 0.07 (WM/CSF regression), -0.43 ± 0.08 (WM/CSF/GS regression) and -0.32 ± 0.13 (aggr. AROMA). Figure 5 (b) shows the opposite effect, depicting the violin plots with *r*-value distributions of the significant voxel population from the “aggressive AROMA” procedure. The *r*-value means and standard deviations at these voxel locations corresponding to each regression procedure are: -0.37 ± 0.14 (no physiologic regression), -0.36 ± 0.13 (WM/CSF regression), -0.38 ± 0.13 (WM/CSF/GS regression) and -0.46 ± 0.06 (aggr. AROMA).

**Figure 5.**
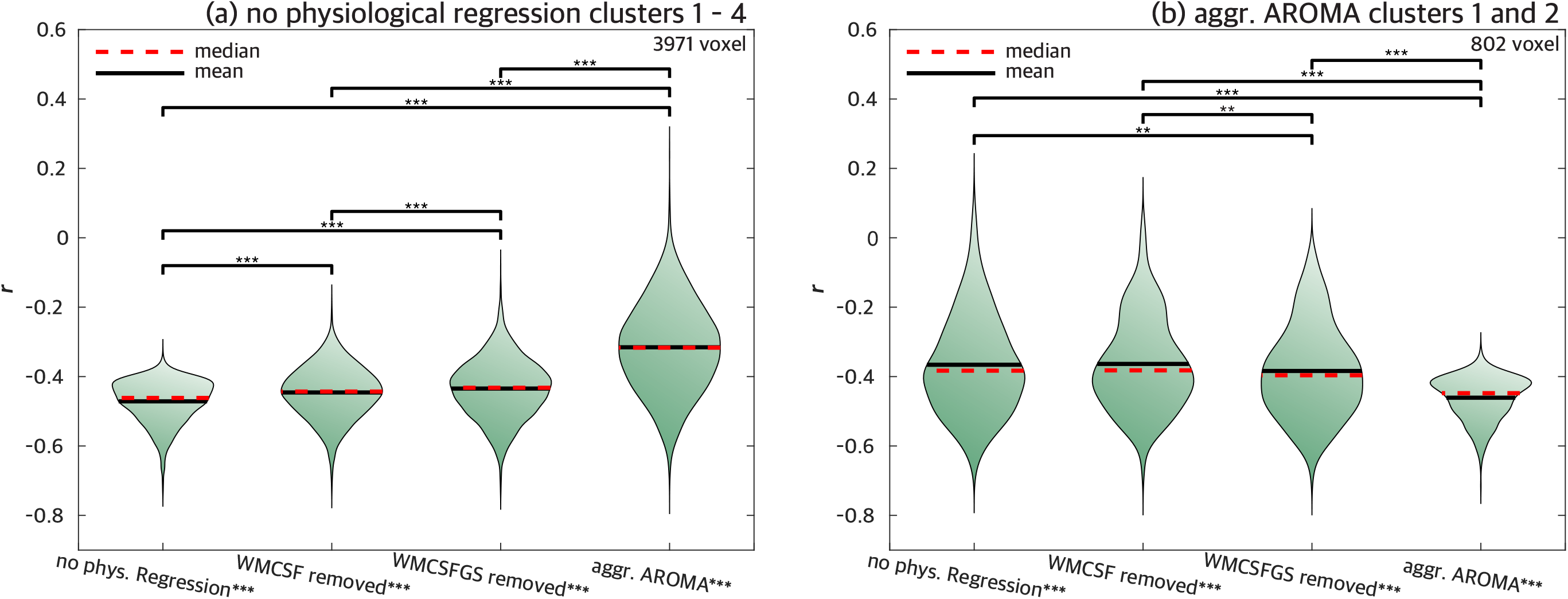
Impact of stepwise regression of upper-stream vascular effects on the correlations between brain amyloid-β deposition and the amplitude of low-frequency BOLD signal fluctuations (sALFF) in the significant clusters identified with “no physiological regression” (a) and “aggressive AROMA” (b). Fisher z transformed r-value distributions shown in violin plots: **(a)** Using the r-values of all voxels within the significant clusters reported in Table 2 under “no physiological regression”. This comparison shows a stepwise decrease of negative correlation magnitude as more aggressive physiological signal removal procedures were applied, suggesting that BOLD-fluctuations at these voxel locations are more driven by upper-stream vascular effects; **(b)** Using the r-values of all voxels within the significant clusters reported in Table 2 under “aggressive AROMA”. This comparison shows a stepwise increase of negative correlation magnitude as more aggressive physiological signal removal procedures were applied, suggesting that BOLD fluctuations at these voxel locations are more likely driven by neuronal activity.

## Discussion

The main finding of this study is the observation of significant negative correlations between the sALFF of rs-fMRI BOLD signals and brain amyloid deposition measured with PET AV45-SUVR at regions associated with default mode and visual networks in patients with amnestic mild cognitive impairment. We have also shown that stepwise removal of the upper-stream physiological/vascular signals reduced the correlation strength between sALFF and AV45-SUVR at these regions. However, even the most intensive upper-stream physiological/vascular signal regression procedures implemented in this study did not completely abolish the negative correlations between sALFF and AV45-SUVR in regions associated with default-mode and visual networks. These findings suggest low-frequency upper-stream cerebral arterial pressure, as well as blood flow fluctuations, are transmitted into the cerebral microcirculation, where they may play an important role in brian Aβ deposition.^33^ Conversely, our data also support the hypothesis that local neuronal activity fluctuations contribute to Aβ homeostasis, independent of the upper-stream vascular effect. Below, we discuss the potential mechanisms and methodological considerations of this study.

### Potential mechanisms

Brain metabolic waste products, including Aβ, are cleared partly via the recently identified brain lymphatic system.^13,62^ This system is a highly organized brain metabolite transportation system between brain interstitial fluid (ISF), cerebrospinal fluid (CSF), and the cerebral vasculature.^13,62^ One theory proposed that CSF enters the perivascular spaces from the subarachnoid space and is propelled deep into the brain by arterial pulsatility. The CSF subsequently enters the neuropil via the astrocytes AQP4 water channels. It then mixes with brain ISF in the extracellular space and leaves the brain, along with brain waste products via the perivenous space into the systemic lymphatic system.^13^ Conversely, other studies have shown that brain waste products also can be cleared via backflow through the periarterial basement space.^62^ Regardless of specific vascular clearance pathways, cerebral arterial pulsatility has been proposed as a key driver of brain waste clearance through these pathways.^14^ Furthermore, a recent study showed that spontaneous cerebral arteriolar vasomotion at a frequency of ∼0.1 Hz was correlated with paravascular clearance of Aβ in the awake mouse brain (van Veluw et al. Neuron 2020), while MRI studies also suggest that low-frequency waves in the CSF of ∼ 0.02 and ∼ 0.05 Hz are likely related to brain Aβ clearance.^63,64^ In this regard, our previous study has demonstrated that brain BOLD signal fluctuations at the regions associated with the default-mode and the visual networks have similar spectral distribution to the fluctuations of systemic blood pressure and CBF velocity measured from the middle cerebral artery, suggesting these upstream vascular signals may transmit downstream into the cerebral microcirculation and impact Aβ clearance.^33^

The present study extended these previous studies by showing that significant negative correlations exist between spontaneous low-frequency BOLD fluctuations and brain amyloid deposition, measured using PET AV45-SUVR, in default-mode and visual networks in patients with amnestic mild cognitive impairment. These findings taken together with the previous studies discussed above suggest that hemodynamic fluctuations transmitted from the upper-stream cardiovascular and cerebrovascular systems into the cerebral microcirculation may influence Aβ homeostasis in individuals with elevated risk for Alzheimer’s disease dementia. Consistent with this hypothesis, we have found that age-related carotid artery stiffness was associated positively with brain Aβ burden in patients with amnestic mild cognitive impairment, suggesting cerebral arterial stiffening may attenuate the transmission of arterial pressure and blood flow oscillations into the brain, leading to a reduction of brain amyloid clearance.^11,65^

Interestingly, even with the most intensive upper-stream physiological/vascular signal removal procedure implemented in this study, negative correlations between sALFF and AV45-SUVR were still present in default-mode and visual regions. These findings suggest that local neuronal activity fluctuations might also prevent brain Aβ deposition.

### Methodological considerations

For calculating the PET AV45-SUVR, we chose white matter instead of the more traditional cerebellum as the reference for normalization. The comparison between reference regions has been comprehensively discussed by Brendel et al..^60^ In their study, they performed a discrimination analysis of healthy controls (HC) vs. mild cognitive impairment vs. Alzheimer’s disease of roughly 1000 subjects from the ADNI dataset. They compared the PET AV45-SUVR discriminative power using different reference regions, including the cerebellum, brainstem, and cortical white matter. Their receiver operating characteristics (ROC) analyses found that using white matter as the reference had the best performance in discriminating the diagnosis groups. Specifically, when compared to the more typical cerebellar reference scaling, a higher discriminatory power between healthy controls and Alzheimer’s disease was found when the white matter was used as a reference. Inter-subject variability was lowest when the white matter was used as a reference as well. Assessment of the longitudinal amyloid deposition was also more reliable and consistent when using white matter as the reference. Using white matter as the reference for the PET AV45-SUVR calculation, Brendel et al. showed that additional partial volume effect correction could lead to 0-2% higher scores in sensitivity and specificity, thus reaching the highest discriminative power of all comparisons. Since the effect of partial volume correction appeared small and its benefits have not been consistent,^66^ we analyzed the AV45 data without partial volume correction. We repeated our analyses with the cerebellum as the reference for AV45-SUVR calculation, finding the same trend of negative correlation between local sALFF and AV45-SUVR (data not shown).

The mean images of the well-established standardized amplitude of low-frequency fluctuations (sALFF) of fMRI recordings in Figures 2 and 3 show a strong effect of preprocessing strategy. As fMRI data is generally suffering from a multitude of confounds and noise signals, the goal of advanced preprocessing strategies is to achieve cleaned BOLD response signals stemming only from neuronal activity. In the present study, we employed different noise regression models, to remove physiological/vascular signals. All regression strategies had inherent motion confound modeling, yet the amount of physiological/vascular regression varied. While white matter (WM) and cerebrospinal fluid (CSF) signals are widely accepted to represent physiological/vascular signal components,^53^ the much-debated global signal has recently been shown to also represent a substantial amount of physiological/vascular signals.^57,67^ Using the data-driven “ICA-AROMA” approach warrants noise-regressors that are closely tailored to the data and promise an effective and robust noise regression,^58^ making it the most demanding regression model in this study.

## Limitations and strengths

Only 33 participants were available in this study and thus the statistical power for voxel-wise analyses was limited. However, our voxel-wise analyses have clearly shown a correlation between sALFF and PET AV45 SUVR. Changes in the correlation strength due to different regression procedures were further presented with cluster-based correlation analyses. However, a causal relationship between brain Aβ deposition and BOLD fluctuation quantified by ALFF cannot be drawn from this cross-sectional study.

The discriminative power of PET AV45 imaging in Alzheimer’s disease has been proven in many studies, nonetheless, there are limitations.^68^ PET images inherently contain a high level of spatial blurring, in addition to a relatively low spatial resolution provided by signal detection. Additionally, the high level of non-specific binding to white matter leads to segmentation and registration inaccuracies at the border between gray- and white matter.^69^ Especially in the situation of grey matter atrophy, commonly observed in Alzheimer’s disease brains, the signal at the cortical grey matter is inevitably contaminated by white matter signal, resulting in reduced specificity. Using the advanced freesurfer segmentation from the anatomical MRI recordings, paired with the manually supervised alignment of AV45 PET images to the anatomical images, we can reduce these segmentation issues in AV45 PET images.

## Conclusions

This study has shown that local spontaneous BOLD signal fluctuations are associated negatively with brain Aβ accumulation measured with amyloid PET in individuals with amnestic mild cognitive impairment who are at a higher risk for Alzheimer’s disease. Furthermore, we found that physiological signal regression decreased the cluster size of negative correlations between low-frequency BOLD signal fluctuation and PET AV45 SUVR. Nevertheless, the most intensive physiological/vascular regression procedures did not completely abolish the correlations between BOLD signal fluctuations and PET AV45 SUVR. Therefore, these findings collectively suggest that both cerebral hemodynamic fluctuations and local neuronal activity play important roles in brain Aβ homeostasis in older adults with amnestic mild cognitive impairment.

## Acknowledgements

We thank each of the study participants for their effort and time contributing to the study. The ^18^F-florbetapir PET radiotracer was provided to the study by Avid Radiopharmaceuticals.

## Funding

Funding towards this work was received from the National Institutes of Health (R01AG033106, R01HL102457, and R01AG057571).

## Competing interests

The authors report no competing interests.

## Supplementary material

Supplementary material is available at *medRxiv* online.

## Abbreviations

ADNI: Alzheimer’s Disease Neuroimaging Initiative
AFNI: Analysis of Functional NeuroImages
ALFF: Amplitude of Low-Frequency Fluctuations
APOE4: Apolipoprotein E4
AROMA: ICA-based Automatic Removal Of Motion Artifacts
AV45: 18F-Florbetapir
Aβ: Amyloid-β
BOLD: Blood-Oxygen-Level-Dependent
CBF: Cerebral Blood Flow
CDR: Clinical Dementia Rating
DPABI: Toolbox for Data Processing & Analysis of Brain Imaging
DPARSFA: Data Processing Assistant for Resting-State fMRI advanced edition
EPI: Echo Planar Imaging
FFT: Fast Fourier Transformation
flirt: FMRIB’s Linear Image Registration Tool
fMRI: functional MRI
FMRIB: Functional Magnetic Resonance Imaging of the Brain
FSL: FMRIB Software Library
FWHM: Full Width at Half Maximum
GS: Global Signal
ICA: Independent Component Analysis
ISF: Interstitial Fluid
MELODIC: Multivariate Exploratory Linear Optimized Decomposition into Independent Components
MMSE: Mini-Mental State Examination
MNI: Montreal Neurological Institute
MPRAGE: Magnetization-Prepared Rapid Acquisition Gradient-Echo
MRI: Magnetic Resonance Imaging
ROC: Receiver Operating Characteristics
rs-fMRI: resting-state fMRI
sALFF: standardized ALFF
SUVR: Standardized Uptake Value Ratio
TE: Time of Echo
TR: Time of Repetition
WM: White Matter
WMS-LM: Wechsler Memory Scale – Revised Logical Memory

## Notes

### Competing Interest Statement

The authors have declared no competing interest.

### Clinical Trial

NCT01146717

### Author Declarations

This study was approved by the Institutional Review Boards of the University of Texas Southwestern Medical Center and Texas Health Presbyterian Hospital of Dallas and performed by the guidelines of the Declaration of Helsinki and Belmont Report.

