## Supplementary material for "Resting-state functional MRI signal fluctuations are correlated with brain amyloid-β deposition"

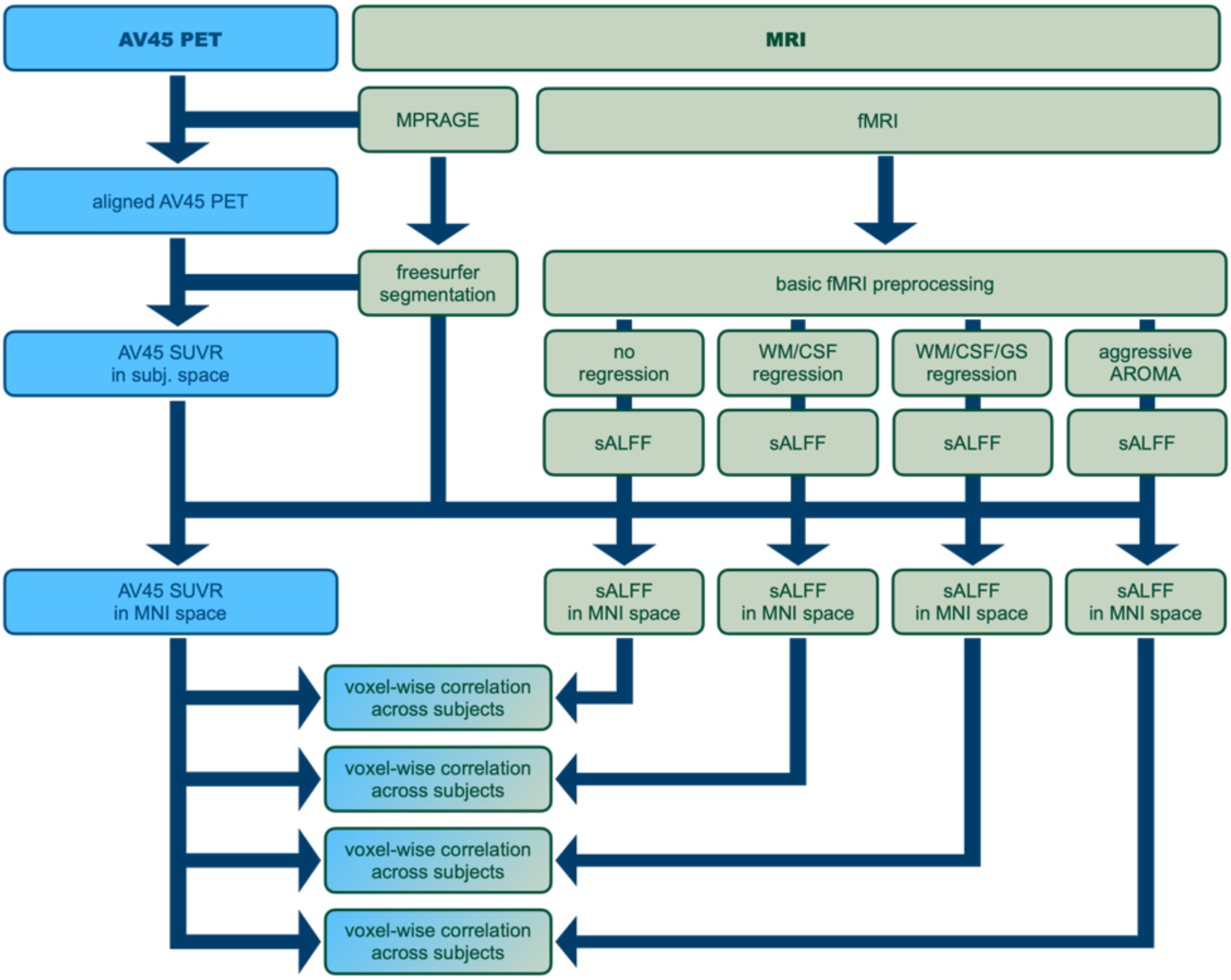


**Sup. Figure 1.** MRI and AV45 PET processing flowchart.

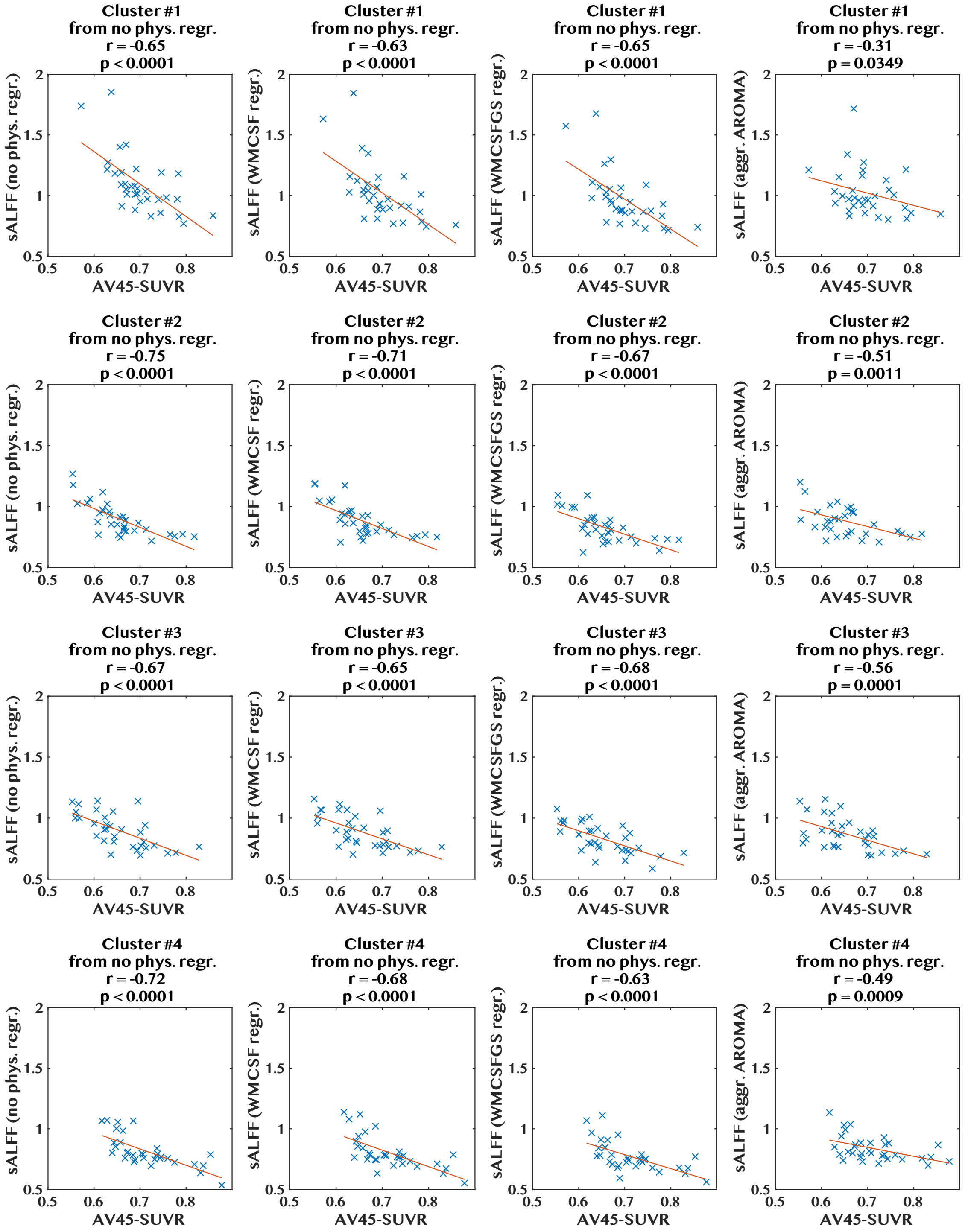


**Sup. Figure 2 Correlation of brain amyloid-β deposition and the amplitude of low-frequency BOLD signal fluctuations (sALFF) - Table 2 "No physiological regression clusters" #1-#4 (rows):** columns depict the scatter-plots for each physiological/vascular regression procedure, left to right: "no physiological regression", "WM/CSF regression", "WM/CSF/GS regression", and "aggressive AROMA". Blue crosses represent the 33 subjects, using mean sALFF and mean AV45 within each cluster. The plots show reduced correlation strength going from column 1 to column 4 for each cluster.

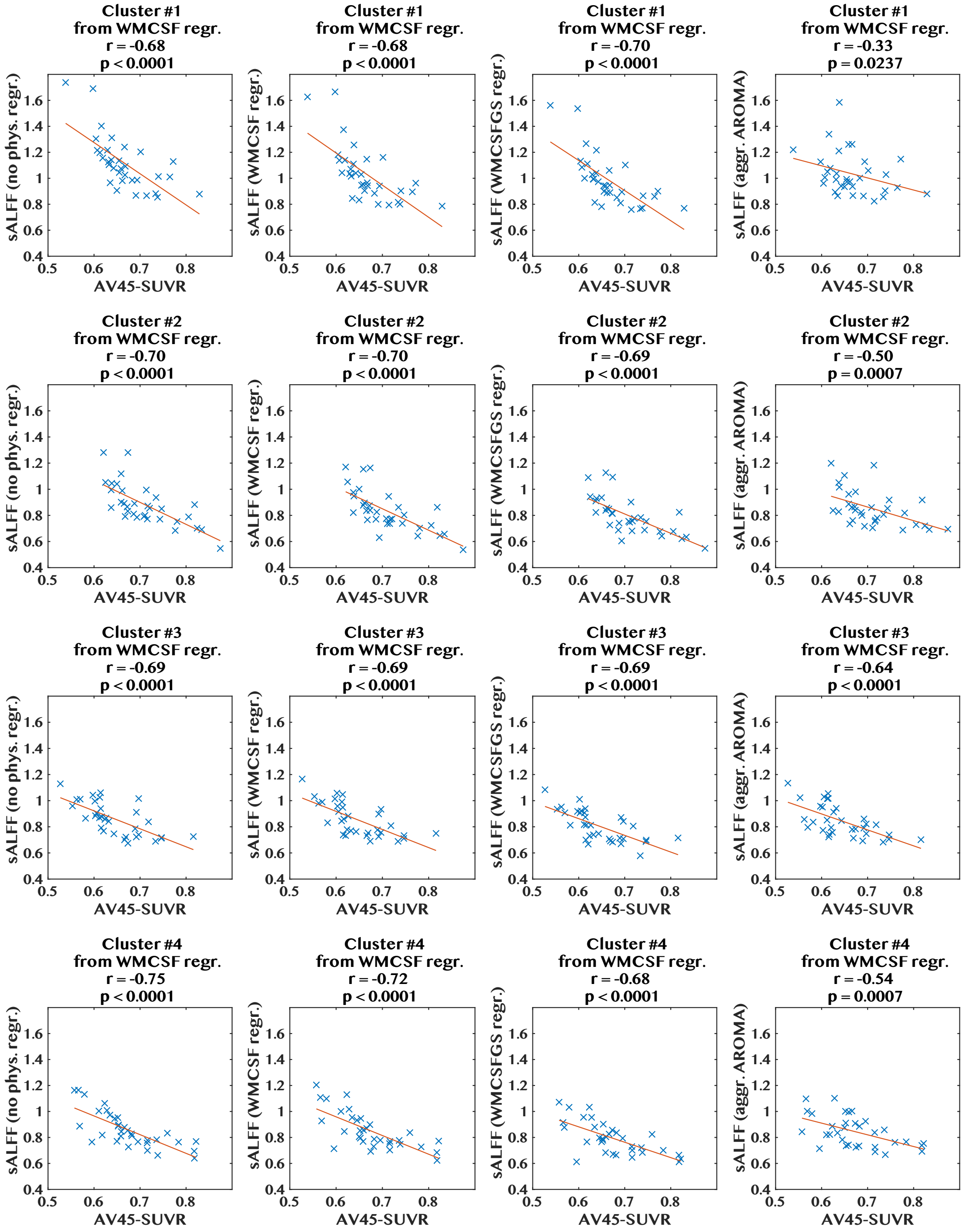


**Sup. Figure 3 Correlation of brain amyloid-β deposition and the amplitude of low-frequency BOLD signal fluctuations (sALFF) - Table 2 "WM/CSF regression clusters" #1-#4 (rows):** columns depict the scatter-plots for each physiological/vascular regression procedure, left to right: "no physiological regression", "WM/CSF regression", "WM/CSF/GS regression", and "aggressive AROMA". Blue crosses represent the 33 subjects, using mean sALFF and mean AV45 within each cluster. The plots show reduced correlation strength going from column 1 to column 4 for each cluster.

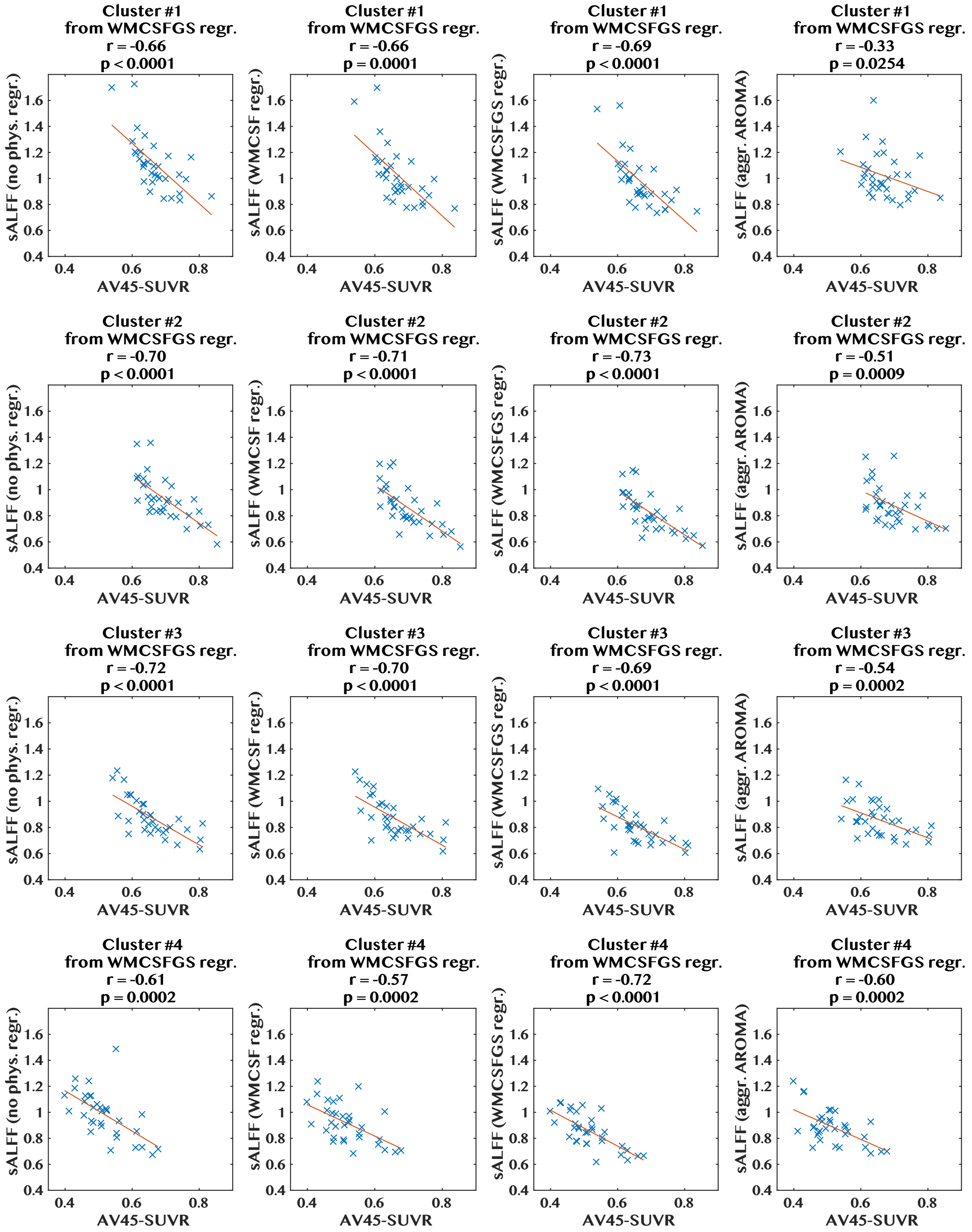


**Sup. Figure 4 Correlation of brain amyloid-β deposition and the amplitude of low-frequency BOLD signal fluctuations (sALFF) - Table 2 "WM/CSF/GS regression clusters" #1-#4 (rows):** columns depict the scatter-plots for each physiological/vascular regression procedure, left to right: "no physiological regression", "WM/CSF regression", "WM/CSF/GS regression", and "aggressive AROMA". Blue crosses represent the 33 subjects, using mean sALFF and mean AV45 within each cluster. The plots show reduced correlation strength going from column 1 to column 4 for each cluster.

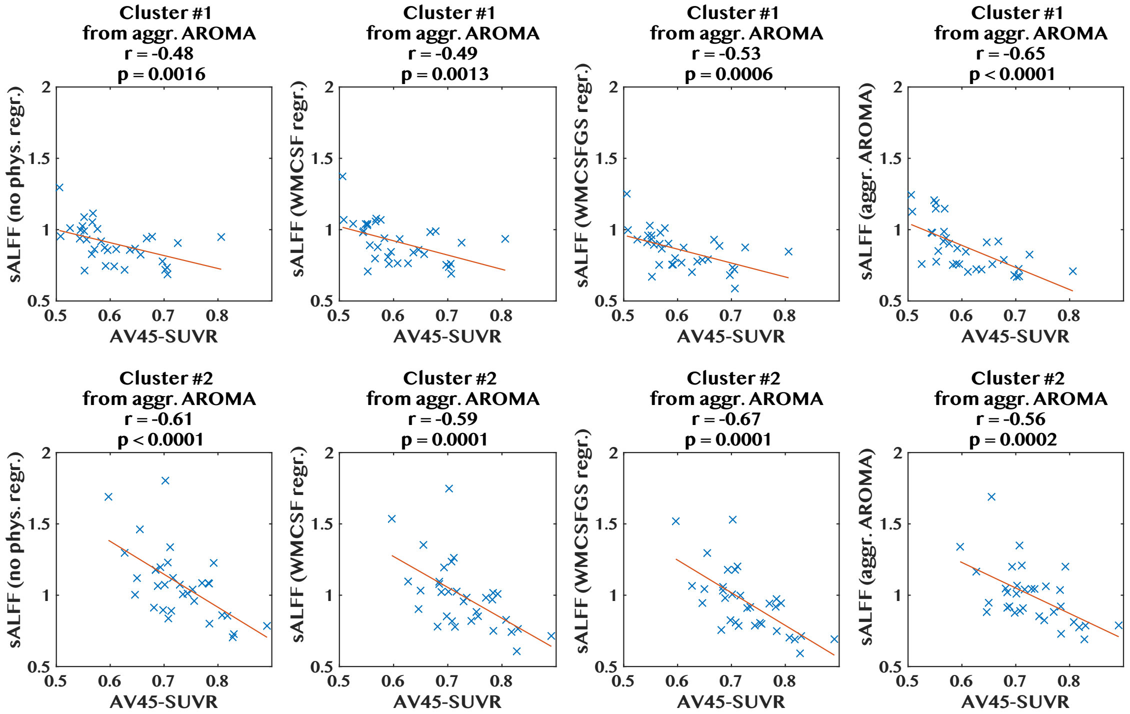


**Sup. Figure 5 Correlation of brain amyloid-β deposition and the amplitude of low-frequency BOLD signal fluctuations (sALFF) - Table 2 "aggressive AROMA clusters" #1 and #2 (rows):** columns depict the scatter-plots for each physiological/vascular regression procedure, left to right: "no physiological regression", "WM/CSF regression", "WM/CSF/GS regression", and "aggressive AROMA". Blue crosses represent the 33 subjects, using mean sALFF and mean AV45 within each cluster. The plot shows increased correlation strength going from column 1 to column 4 for cluster #1.
